# Lack of association between genetic variants at *ACE2* and *TMPRSS2* genes involved in SARS-CoV-2 infection and human quantitative phenotypes

**DOI:** 10.1101/2020.04.22.20074963

**Authors:** Esteban Lopera, Adriaan van der Graaf, Pauline Lanting, Marije van der Geest, Lifelines Cohort Study, Jingyuan Fu, Morris Swertz, Lude Franke, Cisca Wijmenga, Patrick Deelen, Alexandra Zhernakova, Serena Sanna

## Abstract

Coronavirus disease 2019 (COVID-19) shows a wide variation in expression and severity of symptoms, from very mild or no symptomes, to flu-like symptoms, and in more severe cases, to pneumonia, acute respiratory distress syndrome and even death. Large differences in outcome have also been observed between males and females. The causes for this variability are likely to be multifactorial, and to include genetics. The SARS-CoV-2 virus responsible for the infection uses the human receptor angiotensin converting enzyme 2 (*ACE2*) for cell invasion, and the serine protease *TMPRSS2* for S protein priming. Genetic variation in these two genes may thus modulate an individual’s genetic predisposition to infection and virus clearance. While genetic data on COVID-19 patients is being gathered, we carried out a phenome-wide association scan (PheWAS) to investigate the role of these genes in other human phenotypes in the general population. We examined 178 quantitative phenotypes including cytokines and cardio-metabolic biomarkers, as well as 58 medications in 36,339 volunteers from the Lifelines population biobank, in relation to 1,273 genetic variants located in or near *ACE2* and *TMPRSS2*. While none reached our threshold for significance, we observed a suggestive association of polymorphisms within the *ACE2* gene with (1) the use of angiotensin II receptor blockers (ARBs) combination therapies (*p*=5.7×10^−4^), an association that is significantly stronger in females (*p*_*dif*f_=0.01), and (2) with the use of non-steroid anti-inflammatory and antirheumatic products (*p*=5.5×10^−4^). While these associations need to be confirmed in larger sample sizes, they suggest that these variants play a role in diseases such as hypertension and chronic inflammation that are often observed in the more severe COVID-19 cases. Further investigation of these genetic variants in the context of COVID-19 is thus promising for better understanding of disease variability. Full results are available at https://covid19research.nl.

## Introduction

The recent outbreak of the coronavirus disease 2019 (COVID-19) caused by the SARS-CoV-2 virus has quickly become a pandemic and poses a great threat to public health. COVID-19 has a wide range of clinical manifestations: infected people can be asymptomatic, symptomatic with mild respiratory symptoms, or have severe pneumonia (Chen et al. 2020; Wu and McGoogan 2020; Huang et al. 2020; Xu et al. 2020). Estimates based on reported cases from February 2020 in China indicated that approximately 20% of patients develop severe respiratory illness requiring hospitalization, and that overall mortality estimates are around 2.3% (Wu and McGoogan 2020). These estimates are not fixed and are becoming more precise as more cases are reported, screened and analyzed. Interestingly, there is high variability in these estimates when comparing countries and continents, as well as differences in COVID-19 severity between males and females and between different age groups (Wu and McGoogan 2020; Chen et al. 2020) [WHO Situation Report 70, from March 30, 2020]. Differences in response to SARS-CoV-2 infection between individuals and countries may be explained by diminished immune response in the elderly, comorbidities or smoking habits (Guan et al. 2020), but severe COVID-19 cases have also been observed in young individuals, seemingly without risk factors. This indicates that most factors explaining COVID-19 severity are still unknown. It is therefore critical to understand the mechanisms behind COVID-19 severity in order to provide appropriate prevention measures and adequate triage strategies, guide the drug discovery process and ultimately combat the SARS-CoV-2 pandemic.

The large variation in SARS-CoV-2 infection rates and COVID-19 severity could potentially be explained by genetic differences between hosts. While large-scale genetic studies of COVID-19 patients are being assembled, such as those coordinated by the COVID host genetics consortium (https://www.covid19hg.com/), it is worthwhile to evaluate the effects of genetic variants in genes involved in SARS-CoV-2 infection on human phenotypes, including quantitative traits, taking advantage of already existing cohorts. In fact, while quantitative phenotypes are not always directly associated with a disease, knowledge on the genetic variants that modulate these traits can improve our understanding of disease onset and the variability in symptoms. In one example of how this can work, genetic variants in the *BCL11A* gene were associated by genome-wide association studies (GWAS) to fetal hemoglobin (HbF) production in the general population (Menzel et al. 2007), and these genetic variants were subsequently found to modulate the severity of beta-thalassemia and sickle cell diseases (Uda et al. 2008; Lettre et al. 2008). This observation explained why certain individuals were naturally predisposed to mild symptoms of these diseases, while others had very severe clinical outcomes and benefitted from HbF increasing drugs. Therefore, understanding the role of genetic variants at genes essential for SARS-CoV-2 infection in human quantitative phenotypes is important to explain the observed variability in infection susceptibility and severity of COVID-19 and this understanding may suggest potential treatments.

Some factors that are necessary for SARS-CoV-2 infection are known (Hoffmann et al. 2020; Yan et al. 2020). Angiotensin converting enzyme 2 (*ACE2*) is necessary for the invasion of the virus into the host cell through viral spike proteins, and the transmembrane Serine Protease 2 (*TMPRSS2*) is necessary for the correct maturation of these same viral spike proteins that enter the cell through *ACE2* (Yan et al. 2020). According to the GWAS Catalogue (see **URLs**), genetic variants in or near *TMPRSS2*, located on chromosome 21, are associated with susceptibility of prostate cancer and mortality rate in the population, while no associations have been reported for variants in or near *ACE2*. This can be partly explained by the fact that the *ACE2* gene is located on the X chromosome, a part of the genome that is often not analysed by large scale genome wide association studies (GWAS) due to differences in analysis workflow with the autosomal chromosomes. Potential associations with human phenotypes near *ACE2* could have therefore been missed.

Here we investigated the association of genetic variants within or near (±100Kb) *ACE2* and *TMPRSS2* transcripts through a phenome-wide association scan (PheWAS) in 36,339 volunteers from the Lifelines population cohort. We analysed 72 quantitative phenotypes and the use of 58 different drug categories in the entire cohort, and 92 protein levels in plasma and 14 cytokines in a subset of 600 individuals. The quantitative phenotypes selected are anthropometric traits and measurable parameters of lung, hearth, kidney, haematological, immune and cardio-metabolic functions. Finally we evaluated whether these variants differed in their association between males and females to explore potential gender differences that could modulate SARS-CoV-2 infection.

## Materials and Methods

### Lifelines cohort

The Lifelines cohort (Scholtens et al. 2015) is a multi-disciplinary prospective population-based cohort study, with a unique three generation design, that is examining the health and health-related behaviours of 167,729 individuals living in the North of the Netherlands. It was approved by the medical ethics committee of the University Medical Center Groningen and conducted in accordance with Helsinki Declaration Guidelines. All participants signed an informed consent form prior to enrollment. Lifelines employs a broad range of investigative procedures to assess the impact of biomedical, socio-demographic, behavioural, physical and psychological factors on multi-morbidity and complex genetics.

### Genotyping data

A subset of 38,030 volunteers were genotyped using the Infinium Global Screening Array® (GSA) MultiEthnic Disease Version, according to manufacturer’s instructions, at the Rotterdam genotyping center and the Department of Genetics, University Medical Center Groningen. We performed standard quality controls on both samples and markers, including removal of samples and variants with a low genotyping call rate (<99%), variants showing deviation from Hardy-Weinberg equilibrium (p<1×10^−6^) or excess of Mendelian errors in families (>1% of the parent-offspring pairs), and samples with very high or low heterozygosity. We further checked and removed samples that did not show consistent sex information compared to genotypes on the X chromosome, between reported familial information and observed identity-by-descent sharing with family members, and between genotypes available from previous studies(Tigchelaar et al. 2015; Francioli et al. 2015). A detailed description of the process can be found at the following link: https://covid19research.nl (van der Velde et al. 2019). After quality checks, a total of 36,339 samples and 571,420 autosomal and X-chromosome markers were available for analysis.

The genotyping dataset was then imputed using the Haplotype Reference Consortium (HRC) panel v1.1 at the Sanger imputation server (see **URLs**) (Consortium 2015), and variants with an imputation quality score higher than 0.4 for variants with a MAF>0.01 and higher than 0.8 for rare variants (MAF<0.01) were retained. 58.40% of the 21,241 individuals whose genotype passed quality control were female, and the average age at phenotype collection was 39.9 years (±16.3 years).

### Phenotypes

Quantitative phenotypes were measured as previously described (Scholtens et al. 2015). We removed illegal zero or negative values for the ‘QRS’, ‘QT’, ‘HALB’, ‘MAP’, ‘MOP’, ‘EOP’, ‘BAP’, ‘U24HVOL’, ‘ALT’, ‘HR’, ‘EO’, ‘PQ’, ‘MO’ and ‘BA’ phenotypes, and removed -999 values from the electrocardiogram phenotypes ‘P_AXIS’, ‘T_AXIS’ and ‘QRS_AXIS’ (**Supplementary Table 1**). Protein levels in plasma for 92 cardiovascular-related proteins were determined using Olink Proseek Multiplex CVD III panel (OLINK, Uppsala, Sweden), and concentrations of plasma citrulline and cytokines were measured by ProcartaPlex™ multiplex immunoassay (eBioscience, USA) as described previously (Zhernakova et al. 2016; Zhernakova et al. 2018(Zhernakova et al. 2016). Medication use was recorded based on drug packaging brought in by the participant’s on their first visit to the Lifelines inclusion center. Registration of medication use in this way has been shown to be fairly to highly concordant with health record information (Sediq et al. 2018). After conversion to anatomical therapeutic chemical classification (ATC) codes, the first four letters (level 3) were used to define drug categories for association analyses. ATC codes with less than 100 observations were not considered for analysis, leaving 58 drug categories for analysis (**Supplementary Table 2**).

### Statistical Analyses

We tested quantitative phenotypes using linear-mixed models implemented in SAIGEgds v1.0.0 so as to correct for familial relationships and cryptic population structure (Zhou et al. 2018; Zheng et al. 2017). For the X chromosome, genotypes in males were considered diploid. We tested 1,273 genetic variants within and near (±100Kb) *ACE2* (chrX:15,579,156-15,620,271, GRCh37) and *TMPRSS2* (chr21:42,836,478-42,903,043, GRCh37) transcripts. These are all single-nucleotide polymorphisms (SNPs), insertions and deletions with minor allele frequency (MAF) >0.005 that were genotyped or imputed and that passed our quality controls as described above. Analysis through SAIGEgds was carried out for 72 quantitative phenotypes available for all, or a subset of the 36,339 samples (**Table 1**). Drug categories were analysed as binary traits (1=if medication currently in use, 0 otherwise) and restricted only to 1,240 genetic variants with MAF >0.01. In both analyses age and sex were used as covariates. inverse-normal transformation was applied to all quantitative traits prior to model fit. We evaluated gender-specific effects by analyzing males and females separately, using only age as covariate and the same transformations as used for the analysis on the entire cohort.

**Table 1.** Most-significant associations with phenotypes at *ACE2* and *TMPRSS2* loci. The table reports summary statistics for the two most-associated phenotypes and genetic variants at *ACE2* and *TMPRSS2* loci. Positions refer to genome build GRCh 37. Beta indicates the effect for each copy of the alternative allele, in standard deviation units. Ref: reference allele; Alt: alternative allele; AF.Alt: alternate allele frequency; SD: Standard Deviation; EO: Eosinophils; TGL: triglycerides; CHIT1: plasma levels of CHIT1 protein; TR: thrombocytes.

| Trait | Gene | rs.id | Chr:position | Ref/Alt | AF.Alt | Analysis | N | beta (SD) | *p* |
| --- | --- | --- | --- | --- | --- | --- | --- | --- | --- |
| EO | *ACE2* | rs17264937 | X:15647332 | T/C | 0.312 | All | 35494 | 0.416 (0.10) | 1.49x10<sup>-4</sup> |
|  |  |  |  |  |  | Males only | 14,751 | 0.357 (0.146) | 0.0146 |
|  |  |  |  |  |  | Females only | 20,743 | 0.531 (0.159) | 8.17x10<sup>-4</sup> |
| TGL | *ACE2* | rs5980163 | X:15521666 | C/G | 0.016 | All | 36,112 | 0.071 (0.019) | 1.63x10<sup>-4</sup> |
|  |  |  |  |  |  | Males only | 15,004 | 0.108 (0.031) | 4.12x10<sup>-4</sup> |
|  |  |  |  |  |  | Females only | 21,108 | 0.014 (0.021) | 0.488 |
| CHIT1 | *TMPRSS2* | rs150965978 | 21:4294265<br>2 | C/A | 0.063 | All | 526 | -0.630 (0.131) | 2.13x10<sup>-6</sup> |
|  |  |  |  |  |  | Males only | 241 | -0.502 (0.209) | 0.017 |
|  |  |  |  |  |  | Females only | 285 | -0.555 (0.175) | 0.002 |
| TR | *TMPRSS2* | rs28401567 | 21:4295181<br>3 | C/T | 0.166 | All | 36,049 | -2.50 (5.83) | 1.77x10<sup>-5</sup> |
|  |  |  |  |  |  | Males only | 14,975 | -1.82 (0.788) | 0.0210 |
|  |  |  |  |  |  | Females only | 21,074 | -2.74 (0.775) | 4.04x10<sup>-4</sup> |

The 92 circulating plasma proteins and 14 cytokines were measured in a small subset of unrelated individuals and thus did not require correction for familial relationships. These were analysed using PLINK v2.00a3LM. We performed the association mapping with both sexes jointly, or separately as described above, and using inverse-normal transformation on the trait. We analyzed each variant and trait combination with or without the inclusion of age and sex covariates, because some genetic variants combined with the low sample size were too highly correlated with the covariates, making an estimate impossible.

## Results

### Quantitative phenotypes

Using a linear-mixed model, we analyzed 1,273 common and low frequency (minor allele frequency - MAF>0.005) genetic variants in and near (+/-100Kb) *ACE2* and *TMPRSS2* transcripts for association with 178 quantitative traits (**Supplementary Table 1)**. None were found to be significant at the standard genome-wide level (*p*=5×10^−8^) or at the Benjamin-Hochberg false discovery rate (FDR<0.1). The most significant associations found with quantitative traits at the *ACE2* locus were with triglycerides (rs5980163, *p*=1.6×10^−4^) and with the eosinophil counts (rs17264937, *p*=1.5×10^−4^). The strongest associations at the *TMPRSS2* locus were with plasma levels of CHIT1 (rs150965978, *p*=2.1×10^−6^) and thrombocytes (rs28401567, *p*=1.7×10^−5^) (**Table 1**). Only the association at rs5980163 with triglycerides at *ACE2* showed a differential effect between males and females (Cochran Q test *p*_*diff*_=0.01), with the signal observed in the combined analysis with most of the signal being attributable to males, although the association remains only suggestive (*p*=4.12×10^−4^). We did not find any other signal that was restricted to either males or females (*p*<1×10^−6^ for all associations in the gender-specific analyses). When we looked at results obtained for eosinophils, triglycerides and thrombocytes in at least 343,992 samples from the UKBiobank (see **URLs**), none of the associations reported in **Table 1** were found to be significant (all *p*>0.05).

### Medication use

For this analysis, we focused on 1,240 variants with minor allele frequency - MAF>0.01. As with the quantitative phenotypes, none of the genetic variants showed genome-wide significant association with medication use (**Supplementary Table 2**). The strongest associations at the *ACE2* locus were observed for the group of drugs that contains non-steroid anti-inflammatory and antirheumatic products (NSAIDs) (ATC=M01A) (odds ratio (OR)=1.34, 95% C.I.=1.14-1.58, *p*=5.5×10^−4^ for the G allele of rs4646190) (**Table 2**), and for the group that contains angiotensin II receptor blockers (ARBs) in combination with other antihypertensive drugs (ATC=C09D) (OR=1.35, 95% C.I.=1.14-1.62 *p*=5.7×10^−4^ for the T allele of rs4646156) (**Table 2**).

**Table 2.** Top associations with drug prescriptions at *ACE2* and *TMPRSS2* loci. The table reports summary statistics for the two most associated drug categories (given in ATC codes) for genetic variants at *ACE2* and *TMPRSS2* loci. Positions refer to genome build GRCh 37. The Odds Ratio refers to the alternative allele. Ref: reference allele; Alt: alternative allele; OR: Odds ratio; M01A: Anti-inflammatory and antirheumatic products, non-steroids; C09D: Angiotensin II receptor blockers (ARBs), combinations; J02A: Antimycotics for systemic use; D07A: Corticosteroids, plain.

| Drug<br>ATC code | Gene | rs.id | Chr:position | Ref/Alt | AF.Alt | Analysis | N<br>users | OR (95%C.I.) | *p* |
| --- | --- | --- | --- | --- | --- | --- | --- | --- | --- |
| M01A | *ACE2* | rs4646190 | X:15597568 | A/G | 0.0455 | All | 1274 | 1.34 (1.14-1.58) | 5.5x10<sup>-4</sup> |
|  |  |  |  |  |  | Males only | 412 | 1.54 (1.22-1.97) | 3.7x10<sup>-4</sup> |
|  |  |  |  |  |  | Females only | 862 | 1.16 (0.92-1.46) | 0.213 |
| C09D | *ACE2* | rs4646156 | X:15597043 | A/T | 0.646 | All | 202 | 1.36 (1.14-1.62) | 5.71x10<sup>-4</sup> |
|  |  |  |  |  |  | Males only | 88 | 1.14 (0.92-1.42) | 0.231 |
|  |  |  |  |  |  | Females only | 114 | 1.78 (1.35-2.34) | 4.69x10<sup>-5</sup> |
| J02A | *TMPRSS2* | rs457274 | 21:42792485 | C/G | 0.4062 | All | 523 | 1.33 (1.16-1.51) | 3.36x10<sup>-4</sup> |
|  |  |  |  |  |  | Males only | 234 | 1.23 (1.01-1.50) | 0.041 |
|  |  |  |  |  |  | Females only | 289 | 1.41 (1.25-1.60) | 1.46x10<sup>-4</sup> |
| D07A | *TMPRSS2* | rs9975623 | 21:42920296 | A/G | 0.3044 | All | 897 | 1.23 (1.07-1.36) | 1.04x10<sup>-4</sup> |
|  |  |  |  |  |  | Males only | 348 | 1.29 (1.09-1.51) | 2.52x10<sup>-3</sup> |
|  |  |  |  |  |  | Females only | 549 | 1.19 (1.04-1.35) | 0.011 |

NSAIDs are used for treating pain, fever and inflammation, and include ibuprofen. The significance of rs4646190 was stronger in males (*p*=3.7×10^−4^) than in females (*p*=0.08), but the effect sizes were not statistically different (*p*_*diff*_ =0.054).

The second group of drugs encodes for a combined therapy used to treat hypertension. Combination therapy of ARBs with other hypertensive drugs is usually initiated as a second option when the antihypertensive effect of an ARB alone is not sufficient (Flack 2007; Ram 2004). Our results indicate that individuals carrying at least one T allele at the rs4646156 polymorphism were more likely to take this combined therapy compared to individuals with the other allele. The effect of this SNP was also not significant when considering only ARBs intake (ATC=C09C, *p*=0.66). Thus, the association with ARB combination therapies could indicate that individuals in whom it is difficult to manage hypertension may be genetically predisposed to this state by rs4646156. Interestingly, when analysing males and females separately, we found that the effect of rs4646156 on ARB combination therapy was female-specific, even when accounting for differences in number of users (OR=1.78, 95% C.I.=1.35-2.34, *p*=4.7×10^−5^ in females vs OR=1.14, 95% C.I.=0.92-1.42, *p*=0.23 in males, *p*_*diff*_ =0.01).

However, none of these associations meet either the genome-wide or FDR thresholds for significance, so larger sample sizes are necessary to confirm our observations.

The strongest associations in the *TMPRSS2* locus were observed for the group of drugs containing antimycotics prescriptions (ATC=J02A) (*p*=3.65×10^−5^) and for corticosteroids (ATC=D07A) (*p*=1.0×10^−4^). No significant difference with sex was observed for these two associations (*p*_*diff*_ >0.2).

We attempted to validate our findings on medication use using again the UKBiobank public GWAS summary statistics (see **URLs**), although their data refers to the use of individual medications rather than drug categories. When considering the medications in our categories most commonly used (>1,000 users) in the cohort, we found nominal association with the same direction of effects only for glucosamine use (ATC=M01A, *p*=0.002 in the combined analysis and *p*=0.008 in males only) and with candersartan_cilexetil (ATC=C09D, *p*=0.008 in females only) (**Supplementary Table 3**). This lack of replication could be attributable to differences in medication usage reporting between studies. While both are based on self-reported information, in the Lifelines study records are confirmed by medication packaging collected by a nurse during the recruitment.

## Discussion

Recent studies have demonstrated that SARS-CoV-2 uses ACE2 as the key receptor to invade cells (Yan et al. 2020) and that ACE2-mediated cell invasion is enhanced by *TMPRSS2* expression (Hoffmann et al. 2020). Genetic variations in these two genes that interfere with the gene function may thus be involved in the observed variability of SARS-CoV-2 susceptibility and COVID-19 severity. The association of these genetic variants with human phenotypes in the general population may suggest potential treatments and help to better identify at-risk individuals. Here we used a cohort of 36,339 individuals from the Lifelines general population cohort to investigate the impact of variants near and within these two genes on 178 quantitative traits including measurable parameters of lung, hearth, kidney, hematological, immune and cardio-metabolic functions.

We found no significant evidence that common and low frequency variants in these loci were associated with the measured quantitative traits in the general population. We did observe suggestive signals for phenotypes (triglycerides and thrombocytes) that are involved in cardiovascular diseases, which are considered risk factors for COVID-19 diseases (Wu and McGoogan 2020), but none of the genetic variants reached statistical significance despite our large sample size. Nevertheless, we cannot exclude a role of these variants in the regulation of COVID-19 severity through other relevant phenotypes such as specific immune cell types or cytokine levels that were not measured in our cohort.

To evaluate the effect of genetic variation in clinically relevant phenotypes, we investigated the association of genetics with medication use. We observed a marginal association of variants near *ACE2* with use of ARBs combination therapy (ATC=C09D) and with use of non-steroidal anti-inflammatory and antirheumatic drugs (NSAIDs). Interestingly, a marginal association with ARBs (C09C category) was also observed at the *TMPRSS2* locus (rs75833467, p=3.5×10^−4^). These results are intriguing considering the current debate about whether the use of ARBs and NSAIDs could worsen COVID-19 severity (Russell et al. 2020; Little 2020; Kuster et al. 2020), and their potential effect on increasing *ACE2* expression. No significant associations were found for these variants with blood pressure measurements or inflammatory markers in our cohort (p<0.05), not even when the use of such drugs were added as covariates (data not shown). Association with diastolic and systolic blood pressure was also not observed in the large UKBiobank dataset. Thus, these variants are likely to be associated only with clinical conditions such as hypertension and chronic inflammation or with a better drug response.

ARBs are the preferred alternative for patients who experience ACE-inhibitor induced coughing. However, as rs4646156 is not associated with this adverse drug reaction (ADR), our results are likely independent of the switch to ARBs due to ACE-inhibitor induced coughing (Mas et al. 2011). Interestingly, the association of this SNP with ARBs was specific to ARBs combination therapy, thus pointing to individuals with difficult-to-manage hypertension. The major allele (T) of rs4646156 has different frequencies across populations: 0.653 in Europeans, 0.997 in East Asians and 0.797 South Asians, according to 1000 Genomes (see **URLs**). Likewise, the G allele at the rs4646190 SNP, associated with a higher probability of NSAIDs use, shows substantial different frequencies among populations. It is mostly absent in Asians but not in Europeans: 0.03 in Europeans, 0 in East Asians and 0.003 in South Asians, according to 1000 Genomes (see **URLs**).

Through their preferential use of NSAIDS and ARBS combination therapy, the different genotype frequencies in populations and gender-specific effects could partially explain the observed variation in COVID-19 severity. Unfortunately, from our study we cannot prove that these genetic associations are directly related to SARS-CoV-2 susceptibility or COVID-19 severity, nor that ARBs or NSAIDs improve or worsen SARS-CoV-2 susceptibility or COVID-19 severity.

Our study has the following limitations. First, only age and sex were used as covariates in our analyses, which may not be sufficient to correct for confounders for all traits, such as drug usage or diseases, although the effect of these confounders should be mitigated by our large sample size. Secondly, our analyses on medication use are underpowered given the limited number of individuals in the general population who use the medications that we tested, and thus none of the associations found here met the multiple-testing adjusted significance. Third, our results for medication use did not include low frequency and none of the analysis include rare variants (MAF<0.005) which could still be relevant. Fourth, while we can speculate about potential connections of our results with current knowledge of COVID-19, longitudinal and well-characterized data on patients is needed to further explore our hypothesis.

In conclusion we carried out an extensive screening of potential genetic associations at common and low frequency variants in the *ACE2* and *TMPRSS2* genes, and found a lack of substantial effect in human quantitative phenotype variation in the general population. Genetic analyses in more phenotypes are needed to evaluate their functional role in other physiological processes.

Finally, since genetic variation in other genes, for example those involved in regulating the immune system, could also be important in determining SARS-CoV-2 susceptibility and disease severity, large scale genetic initiatives like the COVID-19 host genetics consortium (https://www.covid19hg.com/) that directly involve patients with COVID-19 and deeply characterization of genomes and phenotypes are urgently needed.

### LifeLines Cohort Study - genetic authors

Raul Aguirre-Gamboa (1), Patrick Deelen (1), Lude Franke (1), Jan A Kuivenhoven (2), Esteban A Lopera Maya (1), Ilja M Nolte (3), Serena Sanna (1), Harold Snieder (3), Morris A Swertz (1), Judith M Vonk (3), Cisca Wijmenga (1)

1. *Department of Genetics, University of Groningen, University Medical Center Groningen, The Netherlands*
2. *Department of Pediatrics, University of Groningen, University Medical Center Groningen, The Netherlands*
3. *Department of Epidemiology, University of Groningen, University Medical Center Groningen, The Netherlands*

## Data Availability

The data analysed in this study was obtained from the Lifelines biobank, under project application number OV18_0463. Requests to access this dataset should be directed to Lifelines Research Office.

https://covid19research.nl

## Data availability

Full summary statistics of the results are available at https://covid19research.nl

## Acknowledgements

The authors wish to acknowledge the services of the Lifelines Cohort Study, the contributing research centres delivering data to Lifelines, and all the study participants. We also thank K. McIntyre for editorial assistance, and the UMCG Genomics Coordination center, the UG Center for Information Technology and their sponsors BBMRI-NL & TarGet for storage and computational infrastructure.

This work was supported by the Netherlands Organization for Scientific Research (NWO): NWO Spinoza Prize SPI 92-266 (to C.W.). The Lifelines Biobank initiative has been made possible by funding from the Dutch Ministry of Health, Welfare and Sport; the Dutch Ministry of Economic Affairs; the University Medical Center Groningen (UMCG the Netherlands); the University of Groningen and the Northern Provinces of the Netherlands. The generation and management of GWAS genotype data for the Lifelines Cohort Study is supported by the UMCG Genetics Lifelines Initiative (UGLI). J.F. is supported by NWO Gravitation Netherlands Organ-on-Chip Initiative (024.003.001) and the Netherlands Heart Foundation CVON grant 2018-27. A.Z. is supported by ERC Starting Grant 715772, NWO-VIDI grant 016.178.056, the Netherlands Heart Foundation CVON grant 2018-27, and NWO Gravitation grant ExposomeNL.

## Author Contributions Statement

E.A. and A.v.d.G. performed statistical analyses. E.L., A.vdG., P.L., P.D. A.Z. and S.S. interpreted results. M.v.d.G. and M.S. provided computing infrastructure and web portal; Lifelines Cohort Study, L.F., C.W.. J.F., and A.Z. provided access to the data. E.A., A.v.d.G. And S.S. wrote the manuscript draft with critical input from P.L., L.F., C.W., P.D. and A.Z.. All authors read and approved the manuscript.

## Conflict of interest

The authors declare no conflicts of interest.

## URLs

1000 Genomes study: https://www.internationalgenome.org/

Sanger imputation server: https://imputation.sanger.ac.uk

GWAS catalogue: https://www.ebi.ac.uk/gwas/home (accessed on April 6, 2020)

UK biobank all phenotype associations: http://www.nealelab.is/uk-biobank

## Notes

### Competing Interest Statement

The authors have declared no competing interest.

